# Imputing partial birth dates using day of the week

**DOI:** 10.1101/2025.10.08.25337453

**Authors:** Candice Y. Johnson

## Abstract

**Background:** In deidentified data, exact dates are suppressed to maintain confidentiality of research participants. When the partial date includes only month and year, researchers who need exact dates must impute a day of the month. In some deidentified datasets, day of the week is also provided, but this variable is uncommonly incorporated into the imputation of partial dates. Our objective was to examine the extent to which misclassification is reduced by incorporating day of the week into partial date imputation.

**Methods:** We simulated a population of 594,677 people using the distribution of birthdays in England and Wales in 2024. We imputed birth dates using four methods: (1) first day of the month, (2) 15^th^ of the month, (3) randomly selecting a day of the month, and (4) randomly selecting a day of the month conditional on day of the week. We quantified misclassification as the median number of days between the imputed and true birth date and as the cumulative percentage of the population whose imputed birth date fell within a given number of weeks of their true birth date.

**Results:** Incorporating day of the week reduced misclassification, with a median of 7 days between imputed and exact birth date compared to 8–15 for the other methods. For nearly a quarter of the population, their imputed birth date was their true birth date, compared to 3% in other methods. However, using the 15^th^ day of the month was the best method to ensure that no misclassification was greater than 3 weeks.

**Conclusion:** Incorporating day of the week into random day selection reduced misclassification. This method is easily accomplished in standard statistical software.

## IMPUTING PARTIAL BIRTH DATES USING DAY OF THE WEEK

In deidentified data, exact dates are suppressed to maintain confidentiality of research participants. When the partial date includes only month and year, researchers who need exact dates must impute a day of the month. Selecting an imputation method that minimizes misclassification is important to reduce bias.^1^ Commonly used imputation methods include selecting the first or last day of the month, the 15^th^ day of the month, or randomly selecting a day of the month.^2–4^

In some deidentified datasets, day of the week is also provided because outcomes occur unevenly across the week; examples include U.S. birth and death certificates and hospital discharge data. However, day of the week is uncommonly incorporated into the imputation of partial dates. This omission is a missed opportunity to reduce misclassification because when year and month are known, day of the week narrows the candidate imputed dates from approximately 30 to 5.

Our objective was to examine the extent to which misclassification is reduced by incorporating day of the week into partial date imputation.

We simulated a population of 594,677 people and their true birth dates using the distribution of birthdays in England and Wales in 2024.^5^ We used Stata version 19 (College Station, Texas) for analyses and used its date functions to extract the year, month, day, and day of the week from each true birth date.

Using only year, month, and day of the week, we imputed birth dates using four methods:

(1) first day of the month, (2) 15^th^ of the month, (3) randomly selecting a day of the month, and (4) randomly selecting a day of the month conditional on day of the week. The fourth method involved identifying only the dates that fell on the given day of the week (e.g., Mondays in January 2024) and selecting randomly among those dates.

We quantified misclassification as the median number of days between the imputed and true birth date and as the cumulative percentage of the population whose imputed birth date fell within 0 (no misclassification), 1, 2, 3, 4, or 5 weeks of their true birth date.

Incorporating day of the week reduced misclassification compared to the other methods, with a median of 7 days between imputed and exact birth date compared to 8–15 for the other methods (Table). As expected, by narrowing down the possible dates using day of the week, for nearly a quarter of the population their imputed birth date was their true birth date, compared to 3% in other methods. However, using the 15^th^ day of the month was the best method to ensure that no misclassification was greater than 3 weeks.

**Table.** Median difference between imputed and true birth dates and cumulative percentage of the imputed birth dates falling within a given distance of the true birth dates using four imputation methods.

|  | First Day of<br>Month | 15<sup>th</sup> Day of<br>Month | Randomly<br>Selected Day | Randomly<br>Selected<br>Conditional on<br>Day of Week |
| --- | --- | --- | --- | --- |
| Median difference, days | 15 | 8 | 9 | 7 |
| Cumulative percentage |  |  |  |  |
| 0 days | 3.2 | 3.3 | 3.3 | 23.0 |
| 1 week (1–7 days) | 26.3 | 49.3 | 43.0 | 58.3 |
| 2 weeks (8–14 days) | 49.3 | 95.1 | 72.4 | 82.9 |
| 3 weeks (15–21 days) | 72.4 | 100.0 | 91.3 | 96.8 |
| 4 weeks (22–28 days) | 95.1 | 100.0 | 99.6 | 100.0 |
| 5 weeks (29–30 days) | 100.0 | 100.0 | 100.0 | 100.0 |

Researchers may have good reasons to choose imputation methods that do not incorporate day of the week or minimize misclassification. The first and last days of the month may be desired for start and stop dates to provide conservative estimates of duration. The 15^th^ day may be useful when it is important for all imputed dates to be as close to the true dates as possible but a realistic distribution of dates across the month is not needed.

Randomly selecting a day is a commonly-used method to produce a realistic distribution of dates across the month.^3^ Incorporating day of the week into this random selection reduced misclassification and should be used when day of the week is available.

Incorporating day of the week into the imputation is easily accomplished in standard statistical software. We include example Stata and SAS code in the Supplementary Materials.

## Supporting information

Stata Code

## Data Availability

The data used for this research is available from the UK Office for National Statistics website.

https://www.ons.gov.uk/peoplepopulationandcommunity/birthsdeathsandmarriages/livebirths/bulletins/birthsummarytablesenglandandwales/2024

## REFERENCES

1. Woods LM, Rachet B, Ellis L, Coleman MP. Full dates (day, month, year) should be used in population-based cancer survival studies. International Journal of Cancer. 2012;131(7):E1120–E1124. doi:10.1002/ijc.27545

2. Coles RH, Barnes P, Fingerhut LA, Gentleman JF, Schenker N, Warner M. Imputation of Missing Date Information for Injuries and Poisonings Reported in the National Health Interview Survey. Northeast SAS Users Group 2006. https://www.lexjansen.com/nesug/nesug06/an/da03.pdf

3. Samuels SJ, Cox NJ. Stata Tip 105: Daily Dates with Missing Days. The Stata Journal. 2012;12(1):159–161. doi:10.1177/1536867X1201200110

4. Bowman R. Partial dates; decisions and implications of handling partially missing dates. PHUSE 2006. https://www.lexjansen.com/phuse/2006/po/PO11.pdf

5. Office for National Statistics. Births in England and Wales: Birth Registrations. 2025. https://www.ons.gov.uk/peoplepopulationandcommunity/birthsdeathsandmarriages/livebirths/datasets/birthsinenglandandwalesbirthregistrations

