## Supplementary Materials for "Imputing partial birth dates using day of the week"

### Imputation of Partial Dates Using Day of the Week
#### Supplementary Materials

We use the example of a birth record with the following characteristics to illustrate how to randomly select a day of the month conditional on day of the week:

- Year of birth = 2024
- Month of birth = January
- Day of the week = Tuesday

##### How to conduct the imputation

1. Find the first Tuesday of the month. This is January 2$^{nd}$.
2. Find the last Tuesday of the month. This is January 30$^{th}$.
3. Calculate the number of Tuesdays in the month. This is equal to the number of weeks between the first and last Tuesdays of the month, plus one. There are 5 Tuesdays in January 2024.
4. Randomly select one of those 5 Tuesdays.
5. Use that Tuesday as the imputed date.

##### Sample Stata code

The following 5 lines of code correspond to the 5 steps above.

```
gen firstday = firstweekdayofmonth(month, year, dayofweek)
gen lastday = lastweekdayofmonth(month, year, dayofweek)
gen numberdays = 1 + (lastday - firstday)/7
gen week_random = runiformint(1,numberdays)
gen random_date= firstday + (week_random-1)*7
```

##### Sample SAS code

The following 5 lines of code correspond to the 5 steps above.

```
firstday = nwkdom(1, dayofweek, month, year);
lastday = nwkdom(5, dayofweek, month, year);
numberdays = 1 + (lastday-firstday)/7;
week_random = rand("Integer", 1, numberdays);
random_date = firstday + (week_random-1)*7;
```
