## Supplementary material for "Imputing partial birth dates using day of the week": Stata Code

Statistics/Data analysis

```
/****************************************************************************

BIRTHDATE IMPUTATION

This program imputes day of the month when month, year, and
weekday are available using 4 different methods.

Programmer:             Candice Johnson
Created:                06 OCT 2025

SOURCE OF DATA: https://www.ons.gov.uk/peoplepopulationandcommunity/
birthsdeathsandmarriages/livebirths/datasets/
birthsinenglandandwalesbirthregistrations

****************************************************************************/

**# PREPARE THE DATASET

/*
- Download the dataset (Table 12)
- Delete header rows
- Retain only the variables for date and number of births in 2024
- Rename variables to DATE and BIRTHS
*/


**# DATA CLEANING

*Make all variable names lowercase
rename *, lower

*Create year variable
gen year = 2024
label var year "Year of Birth"

*Create month and day variables
split date, p("/")
rename date1 day
rename date2 month
destring day, replace
destring month, replace
label var day "True Day of Birth"
label var month "Month of Birth"

*Create full date variable
drop date
gen date = mdy(month, day, year)
label var date "True Date of Birth"
format date %td

*Create day of week variable
gen dayofweek = dow(date)
label define dow 0 "0-Sun" 1 "1-Mon" 2 "2-Tue" 3 "3-Wed" 4 "4-Thu" 5 "5-Fri" 6 "6-Sat"
label var dayofweek "Day of Week of Birth"
label values dayofweek dow

*Order variables
order date day month year dayofweek births

*Create one observation per birth and id number
expand births
gen id = _n
label var id "ID"

*Clean up dataset
sort date
order id date day month year dayofweek births
drop births
```

```
**# ASSIGN DATES IN MULTIPLE WAYS

*Set seed
set seed 2069485

*VERSION 1: Assign to first date of month
gen day_first = 1
label var day_first "V1: 1st Day"

*VERSION 2: Assign 15th date of month
gen day_mid = 15
label var day_mid "V2: 15th Day"

*VERSION 3: Assign date randomly
gen day_random = runiformint(1,31) if inlist(month, 1, 3, 5, 7, 8, 10, 12)
replace day_random = runiformint(1,30) if inlist(month, 4, 6, 9, 11)
replace day_random = runiformint(1,29) if inlist(month, 2)

*VERSION 4: Assign randomly conditional on day of week

*Find first of each "day of week"
gen firstday = firstweekdayofmonth(month, year, dayofweek)
format firstday %td

*Find last of each day of week
gen lastday = lastweekdayofmonth(month, year, dayofweek)
format lastday %td

*Find number of each day of week in each month
gen numberdays = 1 + (lastday - firstday)/7

*Assign day to one of the weeks and find corresponding day
gen week_random = runiformint(1,numberdays)
gen day_byday = firstday + (week_random-1)*7
format day_byday %td
drop firstday lastday numberdays week_random


**# QUANTIFY MISCLASSIFICATION

*Numbers of days (absolute) between imputed and true
gen misclass_first = abs(date - mdy(month, day_first, year))
gen misclass_mid = abs(date - mdy(month, day_mid, year))
gen misclass_random = abs(date - mdy(month, day_random, year))
gen misclass_byday = abs(date - day_byday)

*Mean and range of misclassification
tabstat misclass_first, stat(N, mean, p50, min, max)
tabstat misclass_mid, stat(N, mean, p50, min, max)
tabstat misclass_random, stat(N, mean, p50, min, max)
tabstat misclass_byday, stat(N, mean, p50, min, max)

*Categorize misclassification into bins (0, 1-7, etc.)
label define misclass 0 "0 days" 1 "1-7 days" 2 "8-14 days" 3 "15-21 days" 4 "22-28 da
> ys" 5 "29-30 days"

program misclass
        args newvar oldvar
        gen `newvar' = 0 if `oldvar' == 0
        replace `newvar' = 1 if inrange(`oldvar',1,7)
        replace `newvar' = 2 if inrange(`oldvar',8,14)
        replace `newvar' = 3 if inrange(`oldvar',15,21)
        replace `newvar' = 4 if inrange(`oldvar',22,28)
        replace `newvar' = 5 if inrange(`oldvar',29,30)
        label values `newvar' misclass
        tab `newvar', missing
end
```

```
misclass m_first misclass_first
misclass m_mid misclass_mid
misclass m_random misclass_random
misclass m_byday misclass_byday
```
